# Subgingival Microbiome of Type 2 diabetic subjects associated with Periodontal Severity and Metabolic Condition

**DOI:** 10.1101/2025.04.25.25326420

**Authors:** Adriana-Patricia Rodríguez-Hernández, María-de-Lourdes Márquez-Corona, América-Patricia Pontigo-Loyola, Paola Elena García-Vázquez, Christina Murphy, Bruce J. Paster, Laurie-Ann Ximenez-Fyvie

## Abstract

Type 2 diabetes and its risk factors such as dyslipidemia and/or obesity enhance severe periodontitis with a poorly understood microbiome. Hence, the purpose of the present study was to describe and compare the subgingival microbiomes of Type 2 diabetes subjects classified by periodontal diseases, to determine the association between periodontal severity and metabolic condition. 71 DNA subgingival samples of 36 subjects were evaluated by Human Oral Microbiome Identification Microarray using 16S rRNA gene-probe. Blood chemistries were obtained to test hemoglobin, and serum lipid profile. Obesity classification was obtained by Body-Mass-Index. Kruskal-Wallis, Mann-Whitney, Spearman’s correlation, and Multivariate ordinal logistic-regression (periodontal-severity) test were applied (IBM-SPSS-21/Stata 17). Our results suggest samples of subjects with Gingivitis (*n=*12) presented with higher *Bacteroidetes* proportion *vs.* Generalized Periodontitis grouped by severity Stage-I (*n*=17, NS), Stage-II (*n*=30, *p<*0.05), and Stage-III (*n*=12, *p<*0.05); and Generalized Periodontitis Stage-II presented higher *Firmicutes* proportion (*p<*0.05_MW_*vs._*Gingivitis). Human-Microbial-Taxon presented significant higher Score-levels (Levels), frequency (%), or Odds Ratio (OR) of *^1^*Cardiobacterium valvarum*_HMT-540, *Fusobacterium* Cluster-AE01 (Generalized Periodontitis-Stage-I *vs.* Gingivitis_*p<*0.001_Levels, OR: 9.1 and 4.7), *^2^*Porphyromonas gingivalis* HMT-619 (OR: 4.7), ^Ɨ^*Porphyromonas pasteri*_HMT-279 (Gingivitis *vs.* Generalized Periodontitis-Stage-I and II_*p<*0.01/*p<*0.001_Levels), *^3^*Streptococcus constellatus*_HMT-576/*Streptococcus intermedius*_HMT-644 (Generalized Periodontitis-Stages-II, III *vs.* Gingivitis_*p<*0.05_%), and ^†^*Saccharibacteria* (TM7)[G-1]_*bacterium_*HMT-347,350_Gingivitis_*p<*0.05_Levels), some of the Human-Microbial-Taxon identified, presented correlation with total triglycerides (≥150mg/dl, ^Ɨ^0.521), total lipids (≥800mg/dl, ^†^0.478), and obesity (*^1^0.279, *^2^0.280, *^3^0.312). The microbiome of Type 2 diabetes subjects with gingivitis presented the classical microbial profile with representative pathogenic species, while the Generalized Periodontitis Stage I-II subjects presented a microbiome with representative putative and saccharolytic species, some of them strongly associated with the poor control of lipid profile or obesity, and to periodontal-severity.

## Introduction

Diabetes is a major health issue not only because of the increasing number of people suffering from the disease but also because of the limiting complications when people develop diabetes and suffer throughout their lives with chronic metabolic complications [1, 2]. The related morbidity and mortality reflect the high percentage (45%) of undiagnosed people with diabetes, of which type 2 (T2DM) is the most common type. Mexico is the 7^th^ highest number of people worldwide with diabetes (14.1 million) [1, 2].

Several factors may contribute to the presentation of T2DM, and the development of its associated complications such as overweight or obesity, physical inactivity, and poor diet are considered major risk factors [1, 2], which can result in prolonged states of hyperglycemia. Glycated hemoglobin and obesity are currently recognized as the principal cause of a high predisposition to several infections, and it has long been associated with an increased risk of periodontal diseases [3, 4].

Chronic or non-communicable diseases such as cardiovascular diseases, hypertension, or diabetes, plus risk factors such as dyslipidemia or obesity, can develop several macro and microvascular complications [2]. Particularly, T2DM increases oxidative stress, and the advanced glycation end products all together contribute to the severity of periodontal diseases such as periodontitis, characterized by biofilm dysbiosis in susceptible hosts causing loss of tooth-supported tissues and higher severity [3]. Some studies have determined a correlation between destructive periodontal infections and insulin resistance [5]. Evidence supports significant independent associations between periodontal inflammation, glycemic control, and the complications of diabetes. Additionally, there is growing evidence of an independent association between periodontitis and the incidence of diabetes and whereby periodontitis could initiate or aggravate a comorbid diseases including cardio-metabolic, neurodegenerative and autoimmune diseases and cancer [6, 7].

While some studies have suggested that periodontal inflammation and the composition of subgingival plaque can be affected by the degree of glycemic control and/or by the levels of glycated hemoglobin and some dyslipidemia [6]; Furthermore, it remains unclear whether the observed increase in risk and severity of periodontitis in T2DM subjects, is directly related to etiological changes in the microbial composition of subgingival plaque or due to modifications in the inflammatory host-response [8, 9]. Hence, to develop specific and effective treatments over severe periodontal diseases in T2DM individuals, it is important to determine the subgingival microbiome composition and to evaluate the association to endogenous modifiers [5]. There is recent evidence that obesity-related perturbations impact periodontal health and insulin resistance [5]. A more recent study evaluating Mexicans with T2DM presented 3.8 times the probability of presenting severe periodontitis in the presence of dyslipidemia than in individuals without dyslipidemia [5, 10].

Using sequencing molecular techniques of the 16S ribosomal RNA fraction, a high abundance of certain genera has been observed in individuals with T2DM and poor hyperglycemic control, including genders such as *Saccharibacteria* (TM7), *Aggregatibacter*, *Neisseria*, *Gemella, Eikenella* and *Actinomyces* [11–13]. Other reports identified a significant prevalence of saccharolytic species in subjects with poor glycemic control of the families *Streptococcaceae*, *Veillonellaceae* and *Prevotellaceae* [11–14]. Additionally, several studies had reported the low prevalence and bacterial proportion of recognized periodontal pathogenic species such as *Porphyromonas* sp. and *Tannerella* sp. in T2DM individuals with severe periodontitis [11, 13, 15–17]. The question addressed in the present study is whether the subgingival microbiome differs between different periodontal conditions of Mexican diabetes subjects according to the metabolic condition. Therefore, the purpose of the present study was to describe and compare the subgingival microbiomes of T2DM between gingivitis and generalized periodontitis to determine the correlation between periodontal and metabolic condition.

## Methods

### Subject population

The present study was approved by the Ethics Committees of Dentistry School at the Universidad Nacional Autónoma de México and of the of the Health Sciences Institute at Universidad Autónoma del Estado de Hidalgo (ICSA, UAEH) with record numbers UAEH-DI-ICSA-ODO-CF-011 and CEEI-00005-2019. All procedures were explained to participants prior to obtaining their signatures on informed consent forms, confirming their voluntary participation in accordance with the provisions of the Declaration of Helsinki. All research meets the guidelines concerned with biohazardous materials involving all infectious agents or biologically derived infectious materials according to Official Mexican Standard NOM-087-ECOL-SSA1 (2022). Participant recruitment began in February 2013 and concluded in April 2014.

A selected population of 36 subjects diagnosed with T2DM from Hidalgós state, Mexico, were chosen from a cohort transversal study of T2DM individuals who received clinical attention at ICSA, UAEH. Subjects were evaluated by a calibrated clinician with a trial medical history, blood chemistry hemoglobin and lipid profile, and clinical periodontal evaluation.

Inclusion criteria for the selection of the 36 subjects were previous diagnosis of diabetes with < 2 years, with fast glucose > 126 mg/dl., and Mexican descendant by at least two generations and with a complete clinical, periodontal, and microbial trial with at least 20 natural present teeth. Exclusion criteria were current smokers, pregnancy, lactation, antibiotic or anti-inflammatory treatment, smoking, previous periodontal treatment, and no additional systemic diseases and concomitant complications except for diabetes.

Subjects were periodontally classified with Gingivitis (G, *n=*6), and with Generalized Periodontitis (GP, *n=*30). GP subjects were grouped by severity described below, according to the 2017 classification [18]. A total of 71 subgingival samples were analyzed using the Human Oral Microbe Identification Microarray (HOMIM) [19, 20].

### Blood chemistry and clinical evaluation

Blood chemistry elements included fasting glucose to diagnose diabetic individuals, glycated hemoglobin (HbA1c), and to analyze the lipid profile were evaluated total lipids (TL), total triglycerides (TT), total cholesterol (TC), and high and low-density lipids (LDL and HDL). The body mass index (BMI) was obtained for the obesity classification by the formula (kg (weight)/m2 (height)), overweight (BMI: 25.0–29.9), obesity (BMI: 30.0–34.9, class I; 35.0– 39.9, class II; ≥40.0, class III).

### Clinical periodontal evaluation

Periodontal parameters included previous periodontal treatment, number of missing teeth and clinical measurements taken at 6 sites per tooth from all teeth excluding third molars (168 sites per subject), as previously described [21]. The average of the pair of measurements Pocket Depth (PD, mm), Attachment Level (AL, mm), was used in the analyses. Also, dichotomic measurements of plaque accumulation (PLA, 0/1: undetected/detected), gingival erythema (GE, 0/1), Bleeding of Probing (BOP, 0/1), and Suppuration (SUP, 0/1). All parameters were recorded twice by a calibrated examiner using a North Carolina periodontal probe.

Classification of individuals was according to the Consensus report of the workgroup of the 2017 World Workshop between the American Academy of Periodontology (AAP), and the European Federation of Periodontology (EFP), considering the severity of Periodontitis (Stages I-III), the extent and distribution (generalized), and grade (A-C)[18]. Gingivitis subjects had no sites with attachment level loss, in addition to more than 10% to BOP, and no sites with SUP, and additionally were classify as localized (≥ 10% and ≤30% of BOP) or generalized (≥30% of BOP) as a case definition diagnosis [22]. All periodontitis subjects were classified in Generalized Periodontitis with >30% of teeth involved and additionally classified by Severity Stages in Stage I (SI) with interdental AL > 3 mm, Stage II (SII) with AL 3 - 4 mm, and Stage III (SIII) with AL ≥ 5 mm [18]. Stage IV was not considered because of the lack of information about the origin of tooth loss. Grade was defined by Grade B: T2DM subjects with HbA1c <7 and Grade C: T2DM subjects with HbA1c ≥7.0% [18].

### Biofilm subgingival samples

After drying and isolation with cotton rolls, the supragingival plaque was removed with curettes and subgingival plaque samples were obtained with individual sterile 11/12 Gracey curettes (Hu-Friedy) from the distobuccal (DB) site of two selected teeth (excluding third molars) in the follow order 1. Molar (upper and lower) or premolar (upper and lower). All the subgingival samples were provided from two sites with periodontal health (PH, ≤ 2 mm PB) for the Gingivitis subjects, and one with PH and a second one with Periodontitis (Perio, ≥ 3 PB) for the Generalized Periodontitis groups, according to the AL of DB sites of upper or lower 1tst molars.

Samples were placed in individual tubes containing 150 µl of H_2_Odd (Molecular Biology Grade, SIGMA-Aldrich) and placed immediately in Nalgene® Labtop Coolers, –20 °C, Thermo Scientific and transport to start the purification process. Subgingival samples of bacterial DNA were isolated and purified using the DNA Extraction Kit Ready-To-Use (Biosearch Technologies, before Epicentre Kit) with the modify protocol accepted by the Microbial department of The Forsyth institute, Cambridge, M.A., for the microbial identification using by the **H**uman **O**ral **M**icrobe **I**dentification **M**icroarray (HOMIM) [19, 20].

### HOMIM

Sample DNAs were analyzed using the HOMIM surveyed for ∼300 16S rDNA probes to detect the most prevalent Human Microbial Taxon (HMT) [19, 23]. 16S rRNA-based reverse-capture oligonucleotide probes were applied, for each sample, 16S rRNA genes were PCR amplified, and PCR products were then labeled via incorporation of Cy3-dCTP in a second nested PCR as previously reported [22]. The labeled 16S amplicons were hybridized overnight with probes on the slides. After washing, the microarray slides were scanned using an Axon 4000B scanner and crude data was extracted using GenePix Pro software. Data from individual HOMIM signals were translated to a “bar code” format and normalized by comparing individual signal intensities to the average of signals from universal probes, as previews described [24].

### Data statistics

Data statistics included nonparametric Kruskal Wallis (KW), and Mann-Whitney U tests (MW) to compare the groups such as G or GP with SI, SII, and SIII in *phylum* proportion (%), spot levels [1–5] and individual frequency (%) using IBM SPSS-21 statistics software, adjusted for multiple comparisons according to literature [26] with *p*= 0.001274. HMT were grouping by *phylum* as previous described the Human Oral Microbiome Database (HOMD *_v2_*) [24]. The sample calculation was performed for mean differences based on a population of subjects with type 2 diabetes previously evaluated [13], considering a 95% confidence interval (IC), a 5% margin of error (*p<*0.05), and 0.8 as the level of precision, the power calculation yielded a minimum of 44 samples, divided into four groups with a minimum of 11 samples each with.

The Spearman rank correlation coefficient (SC) test was used to determine the associations between all clinical, periodontal, and microbial data with metabolic control. According to the literature [25], the sample power calculation for SC ranked with an IC width at 0.1, sample size requirement between 62 to 1274. In the present study we included 71 samples to calculate 190 variables per analysis using IBM SPSS-21 statistics software using Stata 17 software with IC 95% and Bonferroni correction.

To correlate metabolic values, all T2DM subjects were separate in dichotomic groups (0/1) of fast glucose (<110 and ≥ 110 mg/ 100 mL), HbA1c (< 6.5 and ≥ 6.5 %), TL (<800 and ≥ 800 mg/100 mL), TT (<150 and ≥ 150 mg/100 mL), TC (< 185 mg/ 100 mL and ≥ 185 mg/100), HDL (> 45 mg/10 and ≤ 45 mg/100 mL), LDL (< 100 mg/10 and ≥ 100 mg/100 mL), LDL/HDL radio, < 3 mg/10 and ≥ 3 mg/100 mL), and non-obese and obesity (BMI < 30 and BMI > 30, respectively). SC selected were from 0.1 to < 0.3 with low, from 0.3 to < 0.5 with medium, from 0.5 to < 0.7 with high correlation, and with significant differences with *P* value ≤ 0.05.

Multivariate ordinal logistic regression models were fitted; the dependent variables for each model were the severity of periodontitis (Stages I, II and III). The independent variables included *p* values less than 0.25 in univariate analysis using Stata 17 software with IC 95% and Bonferroni correction.

## Results

The present study included 36 subjects between 10 and 21 months of the previous diagnosis of T2DM and were evaluated clinical periodontal parameters by calibrated clinicians. Subjects’ clinical measurements are described in Table 1. The mean age was 57.6, and a higher percentage of individuals were female (28/36). Both measurements of weight and Body Mass Index (BMI) presented media in the overweight rank. Additionally, the following metabolic parameters were in the rank of poor control: fast glucose (≥ 110 mg/ 100 mL), HbA1c (≥ 6.5 %), TT (≥ 150 mg/100 mL), TC (≥ 185 mg/100), HDL (≤ 45 mg/100 mL), LDL (≥ 100 mg/100 mL), LDL/HDL ratio, ≥ 3 mg/100 mL).

**Table 1.** Clinical and metabolic characteristics of 36 evaluated subjects with T2DM.

| Variable | N= 36 |  |  |  |
| --- | --- | --- | --- | --- |
|  | Media | SEM | Min. | Max. |
| Age (years) | 57.6 | 1.1 | 32.0 | 76.0 |
| Gender (% females) | 77.50 | 0.05 | 0 | 1 |
| Height (cm) | 155.1 | 1.2 | 138.0 | 173.0 |
| Weight (kg) | 66.4 | 1.9 | 48.0 | 114.0 |
| Body Fat (cm) | 29.5 | 1.4 | 16.9 | 60.0 |
| Systolic Blood Pressure (mmHg) | 125.8 | 1.9 | 103.0 | 160.0 |
| Diastolic Blood Pressure (mmHg) | 85.9 | 2.7 | 56.0 | 160.0 |
| Pulse (per minute) | 68.8 | 1.6 | 53.0 | 92.0 |
| Body Mass Index (%) | 27.8 | 0.8 | 18.8 | 46.0 |
| Fast glucose level (mg/100 mL) | 151.3 | 9.4 | 90.0 | 297.7 |
| Glycated hemoglobin A1c (%) | 7.4 | 0.3 | 5.0 | 12.0 |
| Total lipids (mg/100 mL) | 674.5 | 33.5 | 440.0 | 880.0 |
| Total triglycerides (mg/100 mL) | 185.3 | 15.0 | 89.0 | 265.0 |
| Total cholesterol (mg/100 mL) | 195.1 | 10.8 | 138.0 | 274.0 |
| High density lipids (HDL, mg/100 mL) | 40.6 | 1.5 | 32.0 | 55.0 |
| Low density lipids (LDL, mg/100 mL) | 139.6 | 9.1 | 86.0 | 194.0 |
| Atherogenic Index (HDL/ LDL index, %) | 3.6 | 0.3 | 2.0 | 6.0 |

**SEM:** Standard Error of the Mean; **Min.:** Minim; **Max.:** Maxim.

Clinical and metabolic measurements of subjects showed that any variable presented significant differences between periodontal classification (G or GP SI-III) (data not shown).

Subjects were classified periodontally in Gingivitis (*n*= 6), in which 83.3% presented localized one with ≥ 10% and ≤30% of BOP, and 50% of G subjects presented HbA1c ≥6.5% with no evidence of attachment loss with 82.35% of PLA. To classify subjects with GP, they were distributed in 3 groups by stages of severity (GP SI *n=* 9, GP SII *n=* 15, and GP SIII *n=* 6). According to the HbA1c of <6.5 %: Grade B; or ≥6.5 %: Grade C%, the % of each Grade per group were GP SI 75/ 25 %, GP SII 37/ 63 %, and GP SIII 50/ 50 %, B/ C respectively.

The 71 biofilm samples were grouped into G (n= 12), GP SI (17), GP SII (30), and GP SIII (12) shown in Table 2. All the periodontal characteristics were described firstly by the mean of full mouth, in which all variables showed significant differences between groups due to the classification of subjects; and secondary by the periodontal characteristics of the selected teeth of HOMIM test (DB subgingival biofilm samples). All DB sites presented significant differences with higher values of the GP groups, except for PLA, GE, and BOP with no statistical differences after multiple comparisons results. Of 381 probes used in microarray analysis (see Fig S1 Heat map of HOMIM), 301 were positively detected in at least one sample, 167 with ≥ 10% and 134 ≤ 8% of the cumulated %, resulting in a total of 21,371 microbial identifications of the 71 evaluated samples. The *Phylum* proportion of the 301 HMT identified were grouped into the main periodontal groups and GP subjects grouped by *phylum* proportion (see Fig 1).

**Fig 1.**
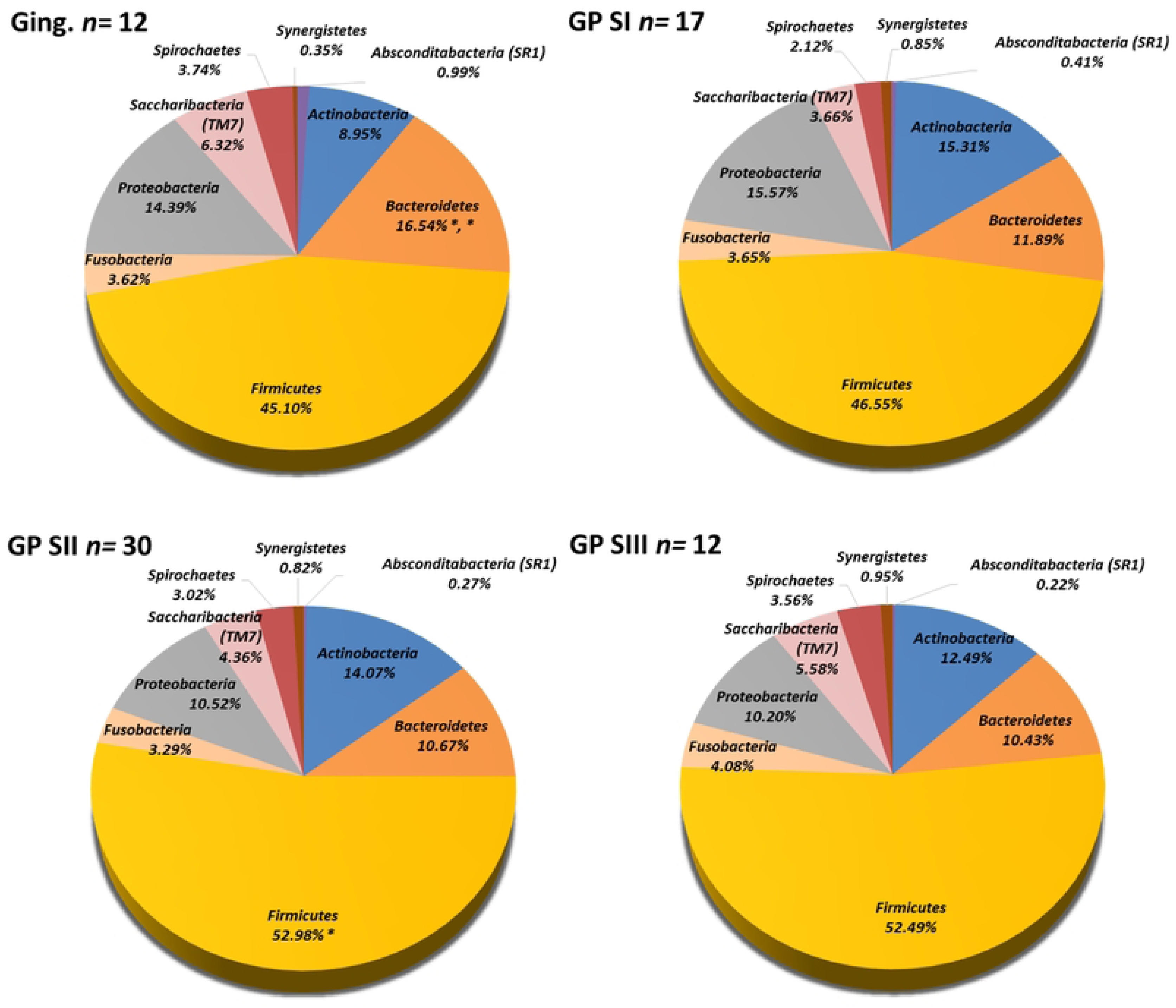
Mean *phylum* proportion (%) of HMT of subgingival biofilm samples from 36 T2DM subjects. Subjects were group into Gingivitis group (*n=* 12 samples from 6 subjects) and Generalized Periodontitis (GP) Stages I-III (SI-SIII, GP SI *n=* 17 samples from 9 subjects, GP SII *n=* 30 samples from 15 subjects, and GP SIII *n=* 12 samples from 6 subjects); The differences were determined between groups of G, GP SI, GP SII and GP SIII with Kruskal Wallis after multiple comparisons correction (Socransky et al., 1991), * *p<*0.05. HMT were grouping by *phylum* as previous described the Human Oral Microbiome Databased (HOMD v2) https://www.homd.org/taxa/tax_table.

**Table 2.**
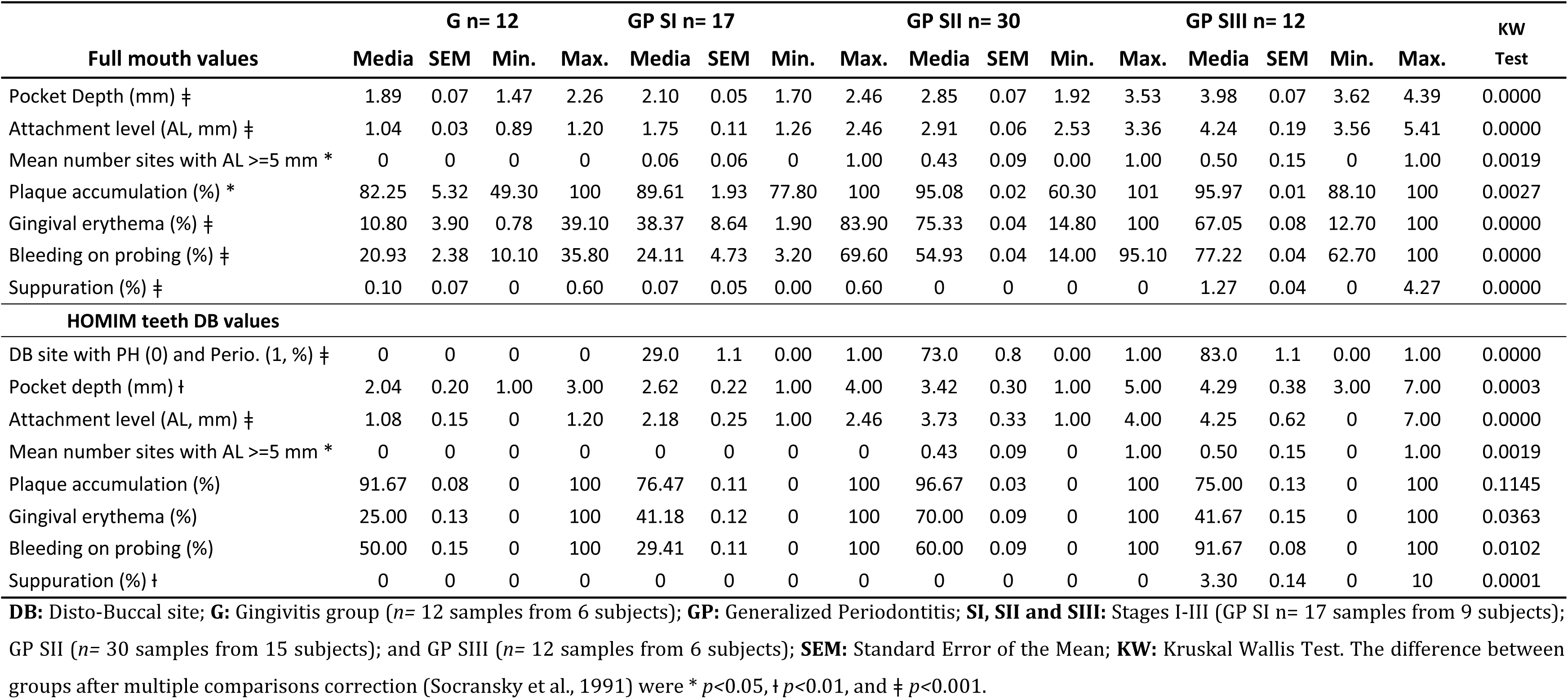
Clinical periodontal characteristics of T2DM subjects with Gingivitis and Generalized Periodontitis Stages I - III.

|  | G n= 12 |  |  |  | GP SI n= 17 |  |  |  | GP SII n= 30 |  |  |  | GP SIII n= 12 |  |  |  | KW |
| --- | --- | --- | --- | --- | --- | --- | --- | --- | --- | --- | --- | --- | --- | --- | --- | --- | --- |
| Full mouth values | Media | SEM | Min. | Max. | Media | SEM | Min. | Max. | Media | SEM | Min. | Max. | Media | SEM | Min. | Max. | Test |
| Pocket Depth (mm) ‡ | 1.89 | 0.07 | 1.47 | 2.26 | 2.10 | 0.05 | 1.70 | 2.46 | 2.85 | 0.07 | 1.92 | 3.53 | 3.98 | 0.07 | 3.62 | 4.39 | 0.0000 |
| Attachment level (AL, mm) ‡ | 1.04 | 0.03 | 0.89 | 1.20 | 1.75 | 0.11 | 1.26 | 2.46 | 2.91 | 0.06 | 2.53 | 3.36 | 4.24 | 0.19 | 3.56 | 5.41 | 0.0000 |
| Mean number sites with AL >=5 mm * | 0 | 0 | 0 | 0 | 0.06 | 0.06 | 0 | 1.00 | 0.43 | 0.09 | 0.00 | 1.00 | 0.50 | 0.15 | 0 | 1.00 | 0.0019 |
| Plaque accumulation (%) * | 82.25 | 5.32 | 49.30 | 100 | 89.61 | 1.93 | 77.80 | 100 | 95.08 | 0.02 | 60.30 | 101 | 95.97 | 0.01 | 88.10 | 100 | 0.0027 |
| Gingival erythema (%) ‡ | 10.80 | 3.90 | 0.78 | 39.10 | 38.37 | 8.64 | 1.90 | 83.90 | 75.33 | 0.04 | 14.80 | 100 | 67.05 | 0.08 | 12.70 | 100 | 0.0000 |
| Bleeding on probing (%) ‡ | 20.93 | 2.38 | 10.10 | 35.80 | 24.11 | 4.73 | 3.20 | 69.60 | 54.93 | 0.04 | 14.00 | 95.10 | 77.22 | 0.04 | 62.70 | 100 | 0.0000 |
| Suppuration (%) ‡ | 0.10 | 0.07 | 0 | 0.60 | 0.07 | 0.05 | 0.00 | 0.60 | 0 | 0 | 0 | 0 | 1.27 | 0.04 | 0 | 4.27 | 0.0000 |
| **HOMIM teeth DB values** |  |  |  |  |  |  |  |  |  |  |  |  |  |  |  |  |  |
| DB site with PH (0) and Perio. (1, %) ‡ | 0 | 0 | 0 | 0 | 29.0 | 1.1 | 0.00 | 1.00 | 73.0 | 0.8 | 0.00 | 1.00 | 83.0 | 1.1 | 0.00 | 1.00 | 0.0000 |
| Pocket depth (mm) † | 2.04 | 0.20 | 1.00 | 3.00 | 2.62 | 0.22 | 1.00 | 4.00 | 3.42 | 0.30 | 1.00 | 5.00 | 4.29 | 0.38 | 3.00 | 7.00 | 0.0003 |
| Attachment level (AL, mm) ‡ | 1.08 | 0.15 | 0 | 1.20 | 2.18 | 0.25 | 1.00 | 2.46 | 3.73 | 0.33 | 1.00 | 4.00 | 4.25 | 0.62 | 0 | 7.00 | 0.0000 |
| Mean number sites with AL >=5 mm * | 0 | 0 | 0 | 0 | 0 | 0 | 0 | 0 | 0.43 | 0.09 | 0 | 1.00 | 0.50 | 0.15 | 0 | 1.00 | 0.0019 |
| Plaque accumulation (%) | 91.67 | 0.08 | 0 | 100 | 76.47 | 0.11 | 0 | 100 | 96.67 | 0.03 | 0 | 100 | 75.00 | 0.13 | 0 | 100 | 0.1145 |
| Gingival erythema (%) | 25.00 | 0.13 | 0 | 100 | 41.18 | 0.12 | 0 | 100 | 70.00 | 0.09 | 0 | 100 | 41.67 | 0.15 | 0 | 100 | 0.0363 |
| Bleeding on probing (%) | 50.00 | 0.15 | 0 | 100 | 29.41 | 0.11 | 0 | 100 | 60.00 | 0.09 | 0 | 100 | 91.67 | 0.08 | 0 | 100 | 0.0102 |
| Suppuration (%) † | 0 | 0 | 0 | 0 | 0 | 0 | 0 | 0 | 0 | 0 | 0 | 0 | 3.30 | 0.14 | 0 | 10 | 0.0001 |

**DB:** Disto-Buccal site; **G:** Gingivitis group ( $n= 12$ samples from 6 subjects); **GP:** Generalized Periodontitis; **SI, SII and SIII:** Stages I-III (GP SI $n= 17$ samples from 9 subjects); GP SII ( $n= 30$ samples from 15 subjects); and GP SIII ( $n= 12$ samples from 6 subjects); **SEM:** Standard Error of the Mean; **KW:** Kruskal Wallis Test. The difference between groups after multiple comparisons correction (Socransky et al., 1991) were \* $p<0.05$ , † $p<0.01$ , and ‡ $p<0.001$ .

*Absconditabacteria* (SR1), and *Bacteroidetes*, showed a higher proportion of G group with no significant differences in Kruskal-Walli’s test. However, the paired comparison of the *phylum* proportion between G, GP SI, GP-II, and GP SIII, *Bacteroidetes* presented higher significant proportions of G group *vs.*GP SII and GP SIII (*p<*0.05), while the *Firmicutes* showed higher proportions of GP SII (*p<*0.05) and GP SIII (NS) *vs.* G group (see Fig 2).

**Fig 2.**
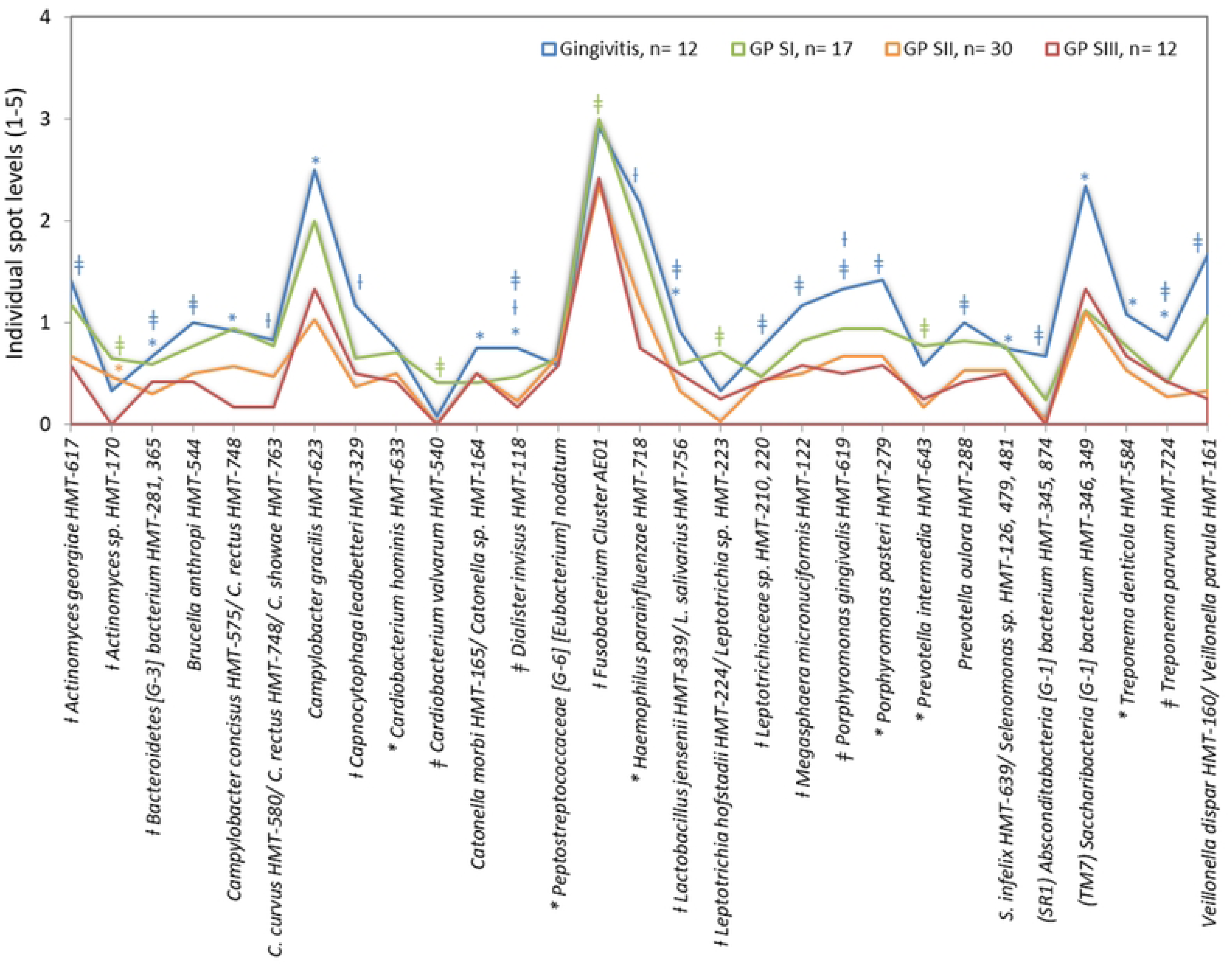
Statistical significance Mean spot levels (1 - 5) of HMT of subgingival biofilm samples from 36 T2DM subjects. Subjects were group into Gingivitis group (*n=* 12 samples from 6 subjects) and Generalized Periodontitis (GP) Stages I-III (SI-SIII, GP SI *n=* 17 samples from 9 subjects, GP SII *n=* 30 samples from 15 subjects, and GP SIII *n=* 12 samples from 6 subjects); The differences were determined between groups of G, GP SI, GP SII and GP SIII groups with Kruskal Wallis (statistics in levels) and Mann Whytney U test (statistics mark in colors by group) after multiple comparisons correction (Socransky et al., 1991), * *p<*0.05 Ɨ *p<*0.01, and ǂ *p<*0.001.

Regarding the relative levels results, after adjusting for multiple comparisons, 29/301 HMT presented significant differences between severity groups with KW and/or MW test (see Fig 2, and Fig. S2). In the paired comparisons. G group presented the highest mean levels of *Catonella morbi*/ *Catonella* sp. (*p<*0.05 G *vs.* GP SI MW); *Absconditabacteria* (SR1) [G-1] *bacterium* (*p<*0.001 G *vs.* GP SII MW); *Actinomyces georgiae*, *Brucella anthropi*, *Leptotrichiaceae* sp., *Megasphaera micronuciformis*, *Porphyromonas pasteri*, *Prevotella oulora* (*p<*0.001 G *vs.* GP SIII MW); *Bacteroidetes* [G-3] *bacterium* (*p<*0.05 G *vs.* GP SII and *p<*0.001 *vs.* GP SIII MW); *Campylobacter concisus*/ *Campylobacter rectus*, *Campylobacter gracilis*, *Saccharibacteria* (TM7) [G-1] *bacterium*, *Treponema denticola* (*p<*0.05 G *vs.* GP SIII MW & KW); *Campylobacter curvus*/ *Campylobacter* rectus/ *Campylobacter* showae, *Capnocytophaga leadbetteri*, *Haemophilus parainfluenzae* (*p<*0.01 G *vs.* GP SIII MW); *Lactobacillus jensenii*/ *Lactobacillus salivarius* (*p<*0.05 G *vs.* GP SI and *p<*0.001 *vs.* GP SIII MW); *Dialister invisus* (*p<*0.05 G *vs.* GP SI, *p<*0.01 *vs.* GP SII and *p<*0.001 *vs.* GP SIII MW); *Porphyromonas gingivalis* (*p<*0.01 G *vs.* GP SI and *p<*0.001 *vs.* GP SII and GP SIII MW); *Veillonella dispar*/ *Veillonella parvula* (*p<*0.001 G *vs.* GP SII and GP SIII MW), *Treponema denticola* (*p<*0.05 G *vs.* GP SI MW); and *Treponema parvum* (*p<*0.05 G *vs.* GP SI and *p<*0.001 GP SIII MW).

In GP SI, *Cardiobacterium valvarum* (KW: *p<*0.001), *Leptotrichia hofstadii*/ *Leptotrichia* sp. (KW: *p<*0.01), *Prevotella intermedia* (KW: *p<*0.05), and *Fusobacterium* Cluster AE01 (*p<*0.001 SI *vs.* G, and *p<*0.01 *vs.* GP SII and SIII MW) were predominant, while *Actinomyces* sp. was predominant in GP SI and SII (*p<*0.001 and *p<*0.05 MW *vs.* G, respectively) (Fig 2, and Fig. S2).

There were statistical differences between groups (% frequency) (Fig 3, and Fig S2). The predominant taxa of the G group were *C. concisus*, *P. pasteri* (*p<*0.05 G *vs.* GP SII MW), and *V. dispar*/ *V. parvula* (*p<*0.01 *p<*0.05 G *vs.* GP SII MW), the predominant frequency taxa of the GP SI were *Veillonella* sp. (*p<*0.01 KW), while the predominant taxa of the GP SII and SIII were *S. constellatus*/ *S. intermedius* (*p<*0.05 GP SII *vs.* G MW), and *Prevotella nigrescens* (*p<*0.05_KW) and *Prevotella pallens* (*p<*0.05_KW) for G and SII. The microbiome of GP SIII represented the simplest microbiome, very similar to GP SII but with 10/ 29 HMT with higher levels than SII in species such as *Bacteroides bacterium* [G-3], *C. gracilis*, and *S. bacterium* (TM7) [G-1] (NS).

**Fig 3.**
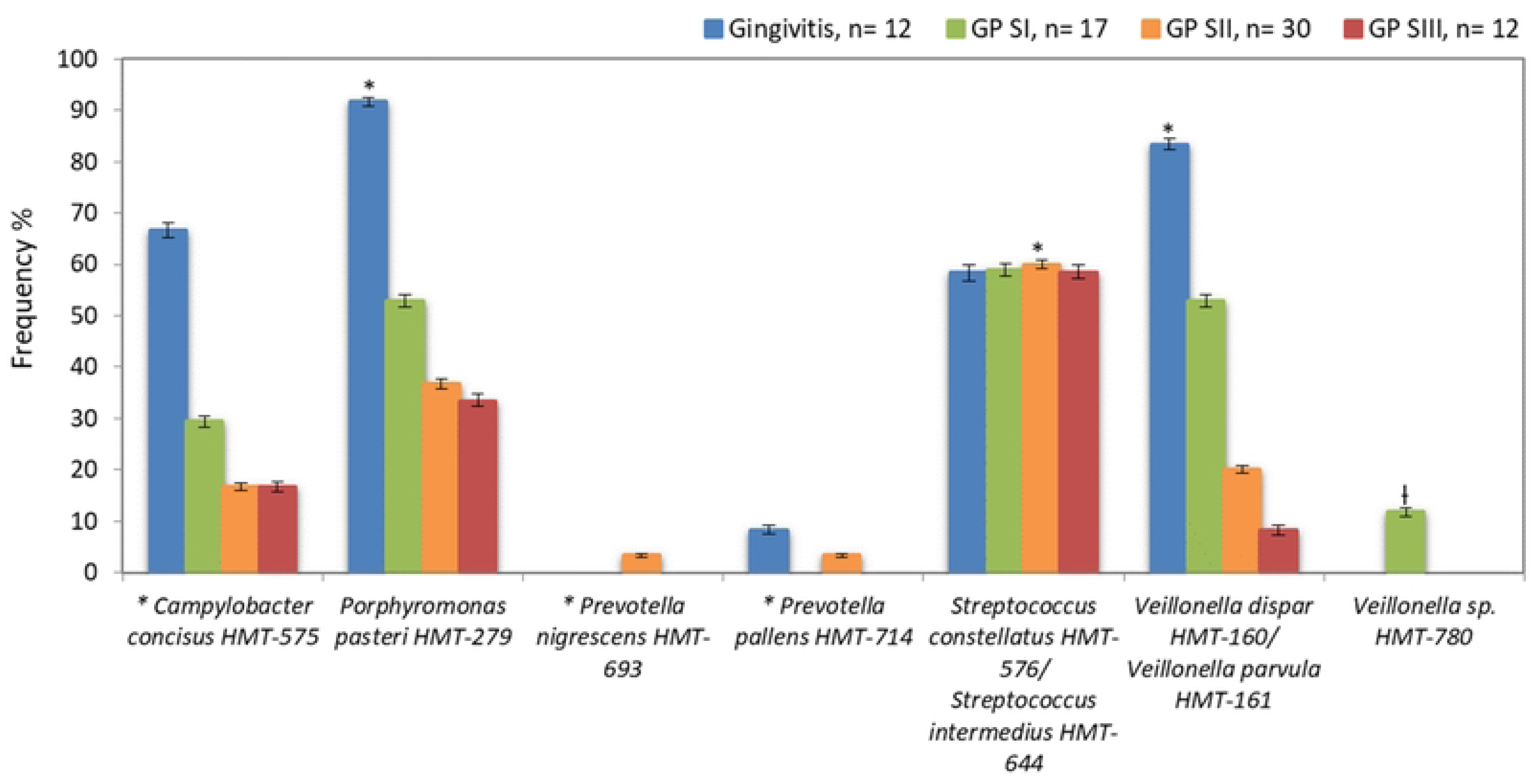
Statistical significance frequency (%) of HMT of subgingival biofilm samples from 36 T2DM subjects. Subjects were group into Gingivitis group (*n=* 12 samples from 6 subjects) and Generalized Periodontitis (GP) Stages I-III (SI-SIII, GP SI *n=* 17 samples from 9 subjects, GP SII *n=* 30 samples from 15 subjects, and GP SIII *n=* 12 samples from 6 subjects); The differences were determined between groups of G, GP SI, GP SII and GP SIII with Kruskal Wallis (statistics in levels) and Mann Whytney U test (statistics mark in colors by group) test after multiple comparisons correction (Socransky et al., 1991), * *p<*0.05 Ɨ *p<*0.01, and ǂ *p<*0.001.

The Spearman correlation tests (see Table 3), presented a positive correlation with significances of measures such as GE (Full mouth) (*p<*0.001) and BOP (Full mouth) (*p<*0.01) with higher and medium correlation of Fast glucose (≥ 150.8 mg/dL), respectively, and with medium correlation of the frequency of *Treponema lecithinolyticum*. GE (Full mouth), the frequency of *Actinomyces* sp. (*p<*0.01), and *Propionibacterium propionicum* (*p<*0.05), presented medium positive correlation with poor control of HgA1c (≥ 6.5 %). Regarding to measurement of total lipids (≥ 800 mg/ dl), *Rothia dentocariosa*/ *Rothia mucilaginosa* frequency (*p<*0.01) and *V. dispar*/ *V. parvula* (*p<*0.001) were highly correlated with total lipids (≥ 800 mg/ 100 mL), as well as *S. bacterium* (TM7) [G-1] (*p<*0.05), presented medium correlation. The measurement of triglycerides (≥ 150 mg/ 100 mL) presented medium correlation with the frequency of *D. invisus*, *H. parainfluenzae* (*p<*0.05), *Lactobacillus gasseri*/ *Lactobacillus johnsonii* and *Selenomonas* sp., (*p<*0.01) while the HMT presented higher correlation were *P. pasteri* and *R. dentocariosa*/ *R. mucilaginosa* (*p<*0.05). Finaly, T2DM subjects with Obesity (I - III), presented medium correlation with the frequency of *Actinomyces gerencseriae*, *Desulfobulbus* sp., *Peptostreptococcaceae* [G-9] [*Eubacterium*] brachy (*p<*0.001), *Fusobacterium naviforme*/ *Fusobacterium nucleatum* subsp. *vincentii* (*p<*0.01), *Helicobacter pylori* (*p<*0.001), *P. gingivalis*, *P. intermedia* (*p<*0.05), *Ruminococcaceae* [G-1] bacterium (*p<*0.01), *Veillonella* sp., (*p<*0.001), and low correlation of *C. valvarum*, and *S. constellatus* (*p<*0.05).

**Table 3.** Associations between clinical, periodontal and microbial data with metabolic control of T2DM subjects.

| Variable | SC | RHO (P value) |
| --- | --- | --- |
| **Fast glucose ($\geq 150.8$ mg/dL)** |  |  |
| Gingival erythema (Full mouth) <sup>H†</sup> | 0.532 | 0.0010 |
| Bleeding on probing (Full mouth) <sup>M†</sup> | 0.467 | 0.0040 |
| *Treponema lecithinolyticum* HMT-653 <sup>M*</sup> | 0.352 | 0.0350 |
| **HgA1c ($\geq 6.5$ %)** |  |  |
| Gingival erythema (Full mouth) <sup>M†</sup> | 0.458 | 0.0050 |
| *Actinomyces* sp. HMT-448 <sup>M†</sup> | 0.457 | 0.0050 |
| *Propionibacterium propionicum* HMT-739 <sup>M*</sup> | 0.331 | 0.0490 |
| **Total lipids ($\geq 800$ mg/ 100 mL)** |  |  |
| *Rothia dentocariosa* HMT-587/ *Rothia mucilaginosa* HMT-681 <sup>H†</sup> | 0.641 | 0.0020 |
| *Saccharibacteria* (TM7) [G-1] *bacterium* HMT-347/HMT_350 <sup>M*</sup> | 0.478 | 0.0330 |
| *Veillonella dispar* HMT-160/ *Veillonella parvula* HMT-161 <sup>H†</sup> | 0.666 | 0.0010 |
| **Triglycerides ($\geq 150$ mg/ 100 mL)** |  |  |
| *Dialister invisus* HMT-118 <sup>M*</sup> | 0.491 | 0.0280 |
| *Haemophilus parainfluenzae* HMT-718 <sup>M*</sup> | 0.448 | 0.0470 |
| *Lactobacillus gasseri* HMT-615/ *Lactobacillus johnsonii* HMT-819 <sup>M†</sup> | 0.360 | 0.0070 |
| *Porphyromonas pasteri* HMT-279 <sup>H*</sup> | 0.521 | 0.0180 |
| *Rothia dentocariosa* HMT-587/ *Rothia mucilaginosa* HMT-681 <sup>H*</sup> | 0.522 | 0.0180 |
| *Selenomonas* sp. HMT-136, 149, 478 <sup>M†</sup> | 0.443 | 0.0010 |
| *Veillonellaceae bacterium* HMT-132 <sup>H*</sup> | 0.553 | 0.0120 |
| **Obesity (I - III)** |  |  |
| *Actinomyces gerencseriae* HMT-618 <sup>M†</sup> | 0.445 | 0.0010 |
| *Cardiobacterium valvarum* HMT-540 <sup>L*</sup> | 0.279 | 0.0430 |
| *Desulfobulbus* sp. HMT-041 <sup>M†</sup> | 0.501 | 0.0000 |
| *Peptostreptococcaceae* [G-9] [*Eubacterium*] *brachy* HMT-557 <sup>M†</sup> | 0.443 | 0.0010 |
| *Fusobacterium naviforme* HMT-689/ *F. nucleatum* subsp. *vinc.* HMT-200 <sup>M†</sup> | 0.405 | 0.0030 |
| *Helicobacter pylori* HMT-812 <sup>M†</sup> | 0.465 | 0.0000 |
| *Porphyromonas gingivalis* HMT-619 <sup>M*</sup> | 0.312 | 0.0230 |
| *Prevotella intermedia* HMT-643 <sup>M*</sup> | 0.304 | 0.0270 |
| *Ruminococcaceae* [G-1] *bacterium* HMT-075 <sup>M†</sup> | 0.407 | 0.0020 |
| *Streptococcus constellatus* HMT-576 <sup>L*</sup> | 0.280 | 0.0430 |
| *Veillonella* sp. HMT-780 <sup>M†</sup> | 0.441 | 0.0010 |

**SC:** Spearman rank correlation coefficient test. **RHO:** P value of Spearman's Correlation with significances represented in labels \* $p < 0.05$ , † $p < 0.01$ , and ‡ $p < 0.001$ . **L, M, and H:** Low, medium and high correlation. **HMT:** Human Microbial Taxon. **HgA1c:** Glycated hemoglobin A1c. ***F. nucleatum* subsp. *vinc.*:** *Fusobacterium nucleatum* subspecies *vincentii*.

The adjusted ordinal logistic regression model showed that in sites with BOP there was a 14% higher risk of developing severe periodontitis and for each year of increase in age, there was a 12% higher risk of developing severe periodontitis. In addition, in samples with higher levels of the pathogenic species such as *C. valvarum* HTM-540 and *P. pasteri* HTM-279, there was a 9.17- and 4.70-times higher risk of developing severe periodontitis, respectively (see table 4). Similar results presented higher proportions of early colonizers of the *phylum Actinobacteria*, *Firmicutes*, there was a 39%- and 42%-higher risk, and of putative and pathogenic species of the *phylum Fusobacteria*, and *Spirochaetes* with 86%- and 68% higher risk of developing severe periodontitis.

**Table 4.** Ordinal logistic regression model of severity of periodontitis (SI-III).

| Variable | Odds Ratio | IC 95% | p |
| --- | --- | --- | --- |
| Body Mass Index | 1.171 | 0.97 – 1.41 | 0.096 |
| **Age (years) \*** | **1.128** | **1.02 – 1.24** | **0.017** |
| Gingival erythema | 0.220 | 0.03- 1.37 | 0.106 |
| **Bleeding on probing ‡** | **1.14e+07** | **750.22 – 1.73e+11** | **0.001** |
| ***Cardiobacterium valvarum* HMT-540 \*** | **9.179** | **1.05 – 80.21** | **0.045** |
| *Treponema lecithinolyticum* HMT-653 | 0.767 | 0.23 - 2.52 | 0.662 |
| *Rothia dentocariosa* HMT-587 | 0.793 | 0.47 - 1.32 | 0.378 |
| *Veillonella dispar* HMT-160 | 0.706 | 0.29 - 1.67 | 0.430 |
| *Dialister invisus* HMT-118 | 0.211 | 0.03 – 1.47 | 0.117 |
| *Haemophilus parainfluenzae* HMT-718 | 0.539 | 0.27 – 1.06 | 0.074 |
| *Lactobacillus gasseri* HMT-615 | 0.224 | 0.02 – 1.90 | 0.171 |
| ***Porphyromonas pasteri* HMT-279 \*** | **4.704** | **1.66 – 18.97** | **0.030** |
| *Selenomonas* sp. HMT-136 | 2.181 | 0.53 – 8.89 | 0.277 |
| *Desulfobulbus* sp. HMT-041 | 0.337 | 0.09 – 1.15 | 0.085 |
| *Absconditabacteria* (SR1) | 0.852 | 0.274 – 2.930 | 0.800 |
| ***Actinobacteria* \*** | **1.391** | **1.000 – 1.936** | **0.050** |
| *Bacteroidetes* | 1.340 | 0.965 – 1.862 | 0.080 |
| ***Firmicutes* \*** | **1.427** | **1.053 – 1.933** | **0.022** |
| ***Fusobacteria* †** | **1.867** | **1.177 – 2.963** | **0.008** |
| *Proteobacteria* | 1.356 | 0.981 – 1.873 | 0.065 |
| *Saccharibacteria* (TM7) | 1.352 | 0.936 – 1.952 | 0.107 |
| ***Spirochaetes* \*** | **1.683** | **1.108 – 2.558** | **0.015** |
| *Synergistetes* | 1.475 | 0.877 – 2.481 | 0.142 |

**p:** P values, Bonferroni adjustment for multiple comparisons according to general characteristics, oral conditions, and mean *phylum* proportion. \* $p<0.05$ , † $p<0.01$ and ‡ $p<0.001$ .

## Discussion

The present study described the microbiome of subgingival biofilm samples of Mexican subjects with diagnosed T2DM, classified with the most recent scheme of periodontal condition classification of the AAP/ EFP [18], even though it was not a healthy group to compare, similar to studies in which the address question was focus of periodontal diseases and metabolic changes of insulin resistance or diabetes individuals [12, 27], we categorized with the metabolic characteristics of T2DM subjects to provide a more comprehensive picture of the association with the etiology and progression of periodontitis still unclear in the literature [7, 8].

HOMIM was very useful in identifying around 300 HMT in the oral cavity and provided a semi-quantification useful to correlate bacterial associations of periodontal [25], even in a small number of samples, thanks to the large number of microbiological identifications as the results in the present study. Although HOMIM is no longer available, we are confident in the HOMIM results as it has been shown that HOMIM and more current NGS technologies show good correlation.[20, 28].

According to diagnostic considerations of gingivitis and periodontitis classification [18], 83.3% of G subjects presented localized gingivitis, and respecting to metabolic data G represented a homogeneous group versus GP-I-III. Some lipid profile values rank in a poor control level, for example, the HDL/LDL ratio denotes a higher level in the evaluated subjects. HDL/LDL ratio is of recent interest to be a dyslipidemia strongly associated with peripheral arterial disease, and consequently to atherosclerosis [29]. In recent studies, the dyslipidemia revealed a significant direct effect and association with inflammation and periodontitis [10, 30], however, no studies relating HDL/LDL ratio with periodontitis severity were shown.

Microbiome results of the T2DM population evaluated in the present study were interesting concerning the description of gingivitis subjects with higher microbial *phylum* proportion and levels than the GP groups. Inflammatory clinical parameters did not present statistical differences between groups, even though the G group had presented higher levels of PLA, GE, and BOP, and additionally, 50% of the G subjects presented HbA1c ≥6.5 %. According to previous studies, Periodontitis severity had been previously associated with HbA1c poor control [31, 32]. However, it is poorly understood how HbA1c levels influence periodontal diseases. A recent study found that gingival inflammation correlated with an HbA1c level of 9.9% of type 1 diabetes [33]. Similar result to in the present study in which GE and BOP correlated individuals with fast glucose (≥ 150.8 mg/dL) and GE with HgA1c (≥ 6.5 %), independently of the periodontal status of T2DM subjects. However, in the logistic regression model of the present study, BOP sites of T2DM subjects presented a 14 higher risk of developing severe periodontitis.

Concerning the microbiome of individuals with diabetes, our results presented a predominance of *Bacteroidetes* in gingivitis subjects with higher levels of some of the HMT such as *C. leadbetteri*, *P. gingivalis*, *P. pasteri*, and *P. oulora*. Members of the *phylum Bacteroidetes* have been previously associated in subjects with T2DM and gingival bleeding suggesting their potential role in the development of diabetes and early stages of periodontal disease [9]. Also, we suggest genders such as *Haemophilus*, *Actinomyces,* or *Veillonella* found with higher proportions in gingivitis individuals of the present study, were more associated with periodontal health conditions such as those recently observed in non-diabetic individuals [34]. On the other hand, in our results the GP SII subjects presented a higher proportion of *Firmicutes* species, in which *Streptococcus* sp. have previously been found in higher levels and proportions in T2DM individuals with chronic forms of periodontitis [13, 35].

A novel result of the present research is the higher presence of *Absconditabacteria* (SR1) with the *A.* (SR1) [G-1] *bacterium* in Gingivitis group. *Absconditabacteria* (SR1) has previously been associated with supragingival biofilm, palatine tonsils, throat, and saliva at the HOMD, however, it is still a gender of a recent study [19]. On the other hand, *D. invisus* and *T. parvum* have been found in higher levels of gingivitis individuals, as shown in our results, which can be a pathogenic niche in T2DM individuals in inflammatory periodontal conditions such as gingivitis. Those periodontal pathogens were recently recognized as novel putative periodontal taxa associated with periodontitis in nondiabetic individuals [36].

Recent studies suggested that further studies were necessary to clarify the effects of hyperglycemia on oral microbial profiles [9, 32]. In the present study, we found the correlation of HMT such as *T. lecithinolyticum* to fast glucose (≥ 150.8 mg/dL) and of *Actinomyces* sp., or *P. propionicum* to HgA1c (≥ 6.5 %). However, it is unclear the periodontal niche of pathogenic taxa in the microbiome of T2DM individuals. One controversial result was the gender *Saccharibacteria* (TM7) previously reported in T2DM subjects with chronic periodontitis and poor glycemic control [11–13]. In the present study, *S. bacterium* (TM7) [G-1] was in higher levels in G individuals, and additionally, it correlated to total poor lipid control. On the other hand, literature suggests that obesity-related perturbations in circulating metabolites may influence the subgingival microbiome [37]. It was evident that the association of putative periodontal pathogens such as *P. gingivalis* correlated to individuals with Obesity (I - III), and the HMT *P. pasteri* correlated to individuals with triglycerides (≥ 150 mg/ 100 mL). In addition, the pathogenic specie *P. pasteri* and *phylum Spirochaetes* presented a higher risk value for severe periodontitis. This suggests that both dyslipidemia of triglycerides and obesity play an important role in the colonization of periodontopathogen bacteria in the T2DM-evaluated individuals with severe periodontitis. The pathophysiological mechanisms, with which the oral microbiota is related to the development of obesity, are not fully understood. In accordance with our results, *P. gingivalis* was detected in higher mean frequency and/ or counts in a Brazilian population of obese women than in non-obese women [38].

Concerning GP SI microbiome, it was represented by putative periodontal species such *P. intermedia* with correlation to Obesity I-III and *Fusobacterium* Cluster AE01 as well as *C. valvarum* correlated with obese individuals, and it was presented in higher risk to severe periodontitis as well as *Fusobacteria phylum* proportion. *C. valvarum* has been identified as the etiology agent of infective endocarditis [39] and was previously negatively associated with individuals with diabetes and periodontitis [31]. However, *Cardiobacterium* sp. has been found in a preview study with significantly differences for gingival index of inflammation in the saliva microbiome of obese children (10-19 years old) than in non-obese and diabetic children [40].

Regarding GP SII and SIII presented a higher mean frequency of nonpathogenic species such as *S. constellatus* correlated to obesity (I-III), *S. intermedius*, and the periodontal putative taxon *P. nigrescens* HMT-693, and concordant with our results, three of the strains were previously found with higher levels or proportions in subgingival microbiota of GP T2DM subjects *vs.* GP non-T2DM [13, 35]. In addition to, T2DM individuals of the present study which presented a higher proportion of *Firmicutes phylum*, they were at risk of presenting severe periodontitis. This *phylum* was previously associated with systemic inflammation of insulin resistance individuals [27].

Finally, the microbiome of GP SIII represented the simplest microbiome of the T2DM evaluated individuals. Compared to the literature, the GP SIII microbial profile of the present study is very similar to a refractory microbial profile, in which the predominance of *Streptococcus* sp. and some putative strains such as *C. gracilis* and low levels of subgingival bacteria [41, 42]. Even though the fact clinical features of refractory periodontitis subjects were different from the T2DM subjects of the present study, host factors can be in common to present a similar microbiome. In the present study, the lowest microbiome levels were more common in the more severe periodontitis of the GP SII and SIII individuals, suggesting an imbalance related to dyslipidemia and obesity presented in T2DM subjects.

## Conclusions

Our findings suggest that poor control of HbA1c (≥ 6.5) was associated only with periodontal inflammation parameters such as GE and BOP but not with a specific T2DM microbiome of the evaluated subjects. Most of the abundant taxa in each evaluated group were associated with at least one metabolic feature such as total lipids, triglycerides, and/or subjects with obesity I-III, and presented higher risk with periodontal severity. This suggests that some dyslipidemia or obesity disturbed the subgingival microbiota of the evaluated subjects more than microbial dysbiosis-promoting by HbA1c.

The GP SI individuals presented a putative pathogenic species such as *C. valvarum* correlated to the obese T2DM of the present study and may serve as a biomarker for cardiovascular disease and severe periodontitis of T2DM individuals, reflected in the subgingival microbiome imbalance.

The microbiome of GP SII and SIII was representing by putative *Firmicutes* species such as *Streptococcus* sp., and the main putative strains still being *Fusobacterium* or *Prevotella* sp., suggesting a microbial dysbiosis of T2DM due to host metabolic characteristics such as obesity or dyslipidemia.

The microbiome of T2DM subjects with Gingivitis presented the classical microbial profile with representative pathogenic species, while GP T2DM (Stage I-II) subjects presented a microbiome with representative putative and saccharolytic species of the nonpathogenic *Firmicutes*, some of them strongly associated with the poor control of the lipid profile or obesity, and to periodontal-severity.

## Data Availability

The data are all contained within the manuscript.

## Acknowledgments

The authors thank the collaboration of the laboratory worker Leticia Cruz Fonseca at UNAM, and the pre-graduated student Jimena Plomares Rodríguez of the Laboratory of Molecular Genetics, School of Dentistry, National Autonomous University of Mexico (UNAM), Mexico City, Mexico, with certification in Quality management systems ISO:9001:2015. Thanks to the support of the scholarship provided by the National Council of Science and Technology (CONACYT): CVU 220510.

## Supporting information

### Supporting Figure legends

**S1 Fig.** Heat map of HOMIM (**H**uman **O**ral **M**icrobe **I**dentification **M**icroarray) of samples from Gingivitis group (*n=* 12 samples from 6 subjects), and Generalized Periodontitis (GP) Stages I-III (SI-SIII, GP SI *n=* 17 samples from 9 subjects, GP SII *n=* 30 samples from 15 subjects, and GP SIII *n=* 12 samples from 6 subjects).

**S2. Fig. A.** Statistical significance Mean spot levels (1 - 5) and **B.** Statistical significance frequency (%) of HMT of subgingival biofilm samples from 36 T2DM subjects. Subjects were group into Gingivitis group (*n=* 12 samples from 6 subjects) and Generalized Periodontitis (GP) Stages I-III (SI-SIII, GP SI *n=* 17 samples from 9 subjects, GP SII *n=* 30 samples from 15 subjects, and GP SIII *n=* 12 samples from 6 subjects); The differences were determined between groups of G, GP SI, GP SII and GP SIII groups with Kruskal Wallis (statistics in levels) and Mann Whytney U test (statistics mark in colors by group) after multiple comparisons correction (Socransky et al., 1991), * *p<*0.05 Ɨ *p<*0.01, and ǂ *p<*0.001.

